# Wastewater-based epidemiology for comprehensive communitywide exposome surveillance: A gradient of metals exposure

**DOI:** 10.1101/2023.09.26.23295844

**Authors:** Lu Cai, Rochelle H. Holm, Donald J. Biddle, Charlie H. Zhang, Daymond Talley, Ted Smith, J. Christopher States

## Abstract

Community wastewater surveillance is an established means to measure health threats. Exposure to toxic metals as one of the key environmental contaminants has been attracting public health attention as exposure can be related to contamination across air, water, and soil as well as associated with individual factors. This research uses Jefferson County, Kentucky, as an urban exposome case study to analyze sub-county metal concentrations in wastewater as a possible indicator of community toxicant exposure risk, and to test the feasibility of using wastewater to identify potential community areas of elevated metals exposure. Variability in wastewater metal concentrations were observed across the county; 19 of the 26 sites had one or more metal results greater than one standard deviation above the mean and were designated areas of concern. Additionally, thirteen of the nineteen sites were of increased concern with levels greater than two standard deviations above the mean. This foundational research found variability in several instances between smaller nested upstream contributing neighborhood sewersheds when measured in the associated downstream treatment plant. Wastewater provides an opportunity to look at integrated toxicology to complement other toxicology data, looking at where people live and what toxicants need to be focused on to protect the health of people in that area.

## 1. Introduction

Wastewater has been used to monitor toxicants including illicit drugs (Banta-Green et al., 2016; Choi et al., 2019; Croft et al., 2020), tobacco metabolites (Choi et al., 2019), and biological agents as weapons (Sinclair et al., 2008). Wastewater surveillance has also been used successfully to monitor disease health threats across geographic scales from buildings and neighborhoods to entire cities (Cohen et al., 2022; Holm et al., 2022; Mercier et al., 2022; National Academies of Sciences, Engineering, and Medicine, 2023; Weidhaas et al., 2021). Exposure to toxic metals as one of the key environmental contaminants has been attracting public health attention as exposure can be related to contamination across air, water, and soil as well as associated with individual factors (Barcelos et al., 2020; Menke et al., 2016; Nadal et al., 2011). Metals voided in human excreta entering a wastewater system can have wide variation; intake of elements is highly dependent on the dietary intake and is equal to the output in human excreta (Rose et al., 2015). However, research into using sub-county wastewater sewersheds to detect smaller neighborhood/multi-neighborhood scales of human exposure to environmental toxicants with spatial variability as part of a comprehensive communitywide exposome surveillance has been limited.

Chronic exposure to environmental pollutants is recognized as even more important than individual genetic predisposition in causing common chronic diseases (Bhatnagar, 2006; Lamas et al., 2023; Vineis et al., 2020). However, clinically relevant toxicity is much less prevalent, and when occurring may present with non-specific symptoms, resulting in a large differential for clinicians to consider (El-Kersh et al., 2022; El-Kersh et al., 2023; Tan et al., 2023).

Metals are categorized based on their various features: transition metals versus non-transition metals based on their chemical features; heavy metals versus non-heavy metals from atomic weight; and essential versus non-essential metals from their biological effects.. Heavy metals are those with either a high atomic weight or density. Essential metals are those required for human health, like zinc and copper; while non-essential metals are not at all necessary for life and include metals such as cadmium, lead, and mercury among others. However, metal toxicity can be caused by both essential and non-essential metals. Essential metals can be toxic when accumulating to excessive levels systemically or in specific tissues or organs. The toxicity caused by nonessential metals can occur directly by the displacement of essential metals in normal biological processes or by binding to reactive groups on biological molecules or indirectly by reducing antioxidant expression and activity thereby dysregulating redox reactions (Briffa et al., 2020; Cai et al., 2005; Zoroddu et al., 2019). Therefore, monitoring metals in drinking water and wastewater is an important surveillance measure for regional public health concerns (Mehnaz et al., 2023; Nabulo et al., 2010; Oloruntoba et al., 2022; States et al., 1985). As well, the difference between drinking water and wastewater concentrations could be an indicator of community metal exposure, once ruling out industrial/commercial discharge, and when compared to national drinking water guidelines (EPA, 2009). That 90% of the American public support wastewater monitoring of toxicants as a standard complement of public health tools indicates the potential for the growth of this form of community health surveillance (LaJoie et al., 2023).

Sewers offer an alternative community health monitoring tool to complement clinical toxicity presentation. Approximately 95% of urban household in the U.S. uses a wastewater treatment sanitation system that could be monitored at a community level (WHO/UNICEF, 2023). The potential for rural community wastewater monitoring via sewered or non-sewered sanitation system also remains (Holm et al., 2023a). This research aims to use Jefferson County, Kentucky, as an urban exposome case study to analyze the sub-county metal concentrations in the wastewater as a gradient of community toxicant exposure, and to test the feasibility of using wastewater to identify community areas of elevated metals exposure.

## 2. Methods

### 2.1 Study site

Our study took place in Louisville/Jefferson County, Kentucky (USA) (Fig. 1). Twenty-six sample locations were selected across the county to represent geographic and demographic sub-county variability, and purposive site selection for neighborhoods with known environmental exposures and risks (Fig. 1). Of the sample locations, 21 were upstream corresponding (sub-county nested area) sewersheds which eventually flowed to a water quality treatment center (WQTC). Some sample locations were a combined sanitary sewer and storm water system, which may cause dilution or contribute toxicants from the environment during high rainfall events. The five downstream WQTCs offer an aggregated 97% coverage of county households (Holm et al., 2022). We assume metals emitted by environmental polluting sources (e.g., Toxics Release Inventory (TRI) facilities) could reach wastewater via disposal of human excreta after human exposure to abovementioned metals, or from environmental polluting facilities in the form of industrial discharge to the sewer through permitted discharge limits or environmentally by wind, rain or surface runoff.

**Fig. 1.**
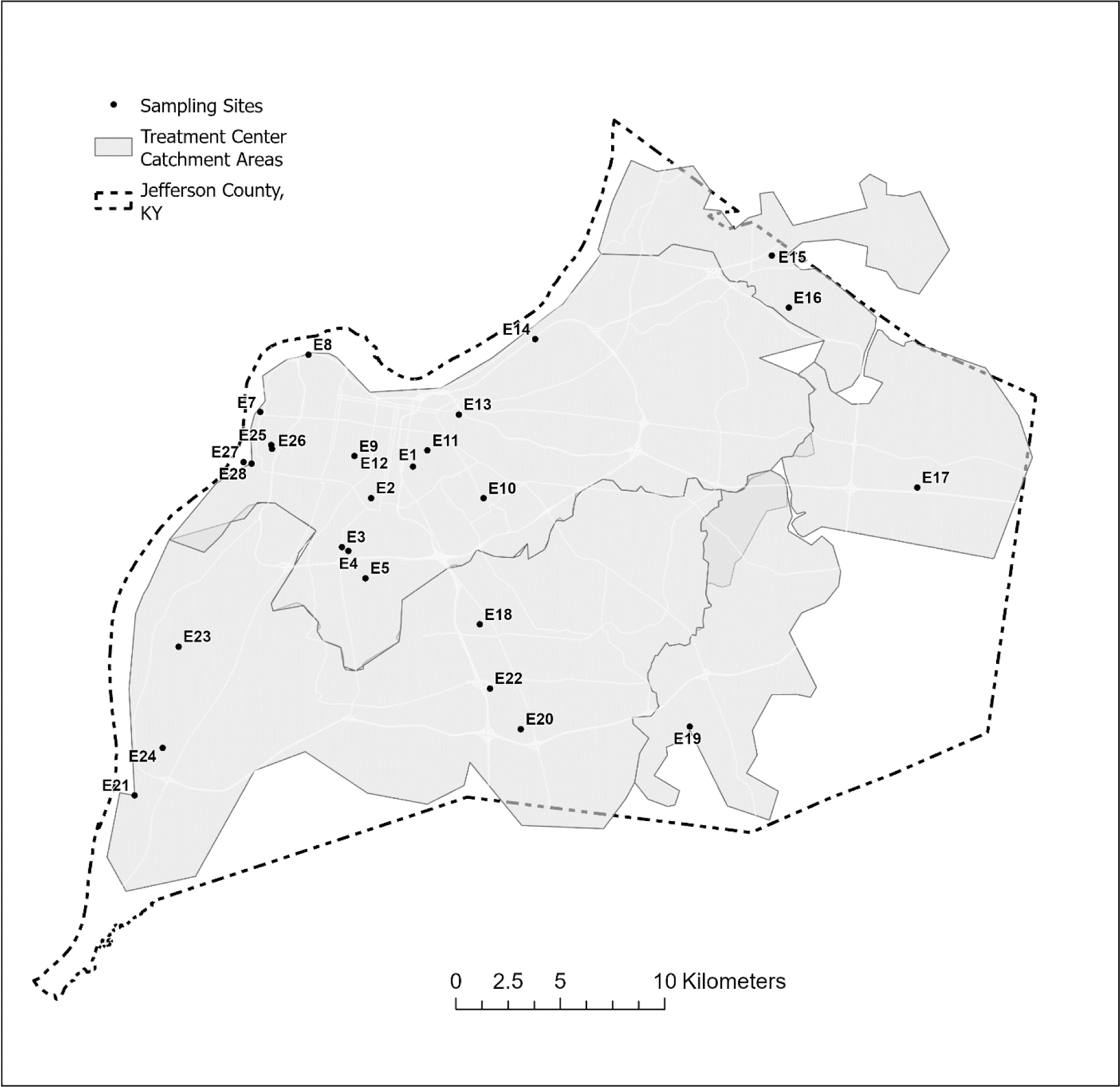
Location of wastewater sampling sites studied, Jefferson County, Kentucky (USA).

### 2.2 Wastewater sample collection and handling

Influent wastewater samples were collected on 12/12/2022 (N = 27). A grab sample was collected into four 50 ml polypropylene tubes per site. Samples were transported on ice to a laboratory in the University of Louisville for analysis. One blank sample using PFAS free water was collected by pouring over the sampling equipment to test for quality assurance and metals contamination in the sample collection process. Additionally, one sample location was sampled at two different times on the same day to test for sample site variability.

### 2.3 Metal analysis

Wastewater samples were stored at -20°C until analysis. Samples were thawed, mixed, 1mL was transferred to a 15 mL metal-free tube (VWR #89049-172) and 3 mL 70% nitric acid (trace metal grade, Fisher Scientific Cat#A509-P500) was added to the sample tube and mixed again. Samples were digested in a 65°C shaker for 5 h until the solution became clear with no residues. After digestion, samples were cooled to room temperature, then diluted 10-fold with deionized water (Millipore, Milli Q Academic). The resulting assay volume was 10 mL. Metal content was measured using an Agilent 7800 ICP-MS (Inductively Coupled Plasma Quadrupole Mass Spectrometer, Agilent Technologies, Japan). Optimization was by performance check with 1 ppb tuning solution, and the assay program was auto tuned by a 10 ppb tuning solution (Agilent Cat#5188-6564). The auto sampler SPS 4 was used for sample introduction. The analysis was performed to test 26 metals. Platinum standard solution (Cat#: CGPTN1) and the calibration standard for the remaining 25 metals (Cat# IV-STOCK-50) were purchased from Inorganic Ventures (Christiansburg, VA). Serial metal standard dilutions were made with the same acid matrix as samples. The internal standard (Cat#5188-6525) was purchased from Agilent. The assay program was run by Agilent MassHunter software with He mode, each sample was read in triplicate and averaged for a final mean value (Table S1).

### 2.4 Data analysis

The Louisville-Jefferson County Metropolitan Sewer District (MSD) provided GIS feature data representing the local sewer system in ESRI Geodatabase format (MSD, 2022). These data included features and attributes for sewer main lines, utility access hole points, and property service connection (PSC) points. Land parcel polygons published by the Jefferson County Property Valuation Administration (PVA, 2022) were also obtained as an ArcGIS Online feature service. A trace network was created from the sewer line features facilitating the use of the Upstream Trace function to select the lines associated with each sampling site. To determine the properties connected to the selected upstream lines, a related feature class of PSC point features was queried. This step allowed us to identify the properties that were directly linked to the sewer infrastructure lines contributing to the sampling location. A location query was then used to select the land parcels containing these service connections, establishing the spatial extent of the properties connected to the selected sewer mains. The selected land parcels, represented as polygons, were converted to point geometry and then aggregated to obtain a single bounding area, effectively delineating the sewershed area for each sampling location. Limited manual feature edits were made to the resulting sewershed area polygons to remove any artifacts from the automated processing. Estimates of selected population variables within each sewershed area were obtained from the U.S Census Bureau using the Enrich tool in ArcGIS Pro (Table 1). Vulnerable communities were determined by overlaying sampled sewersheds with ancillary data on household income and known environmental pollution sources (i.e., TRI facilities, brownfields, superfund sites, and major roads) to explore the spatial associations of metal concentrations in wastewater and aforementioned environmental risk factors.

**Table 1.**
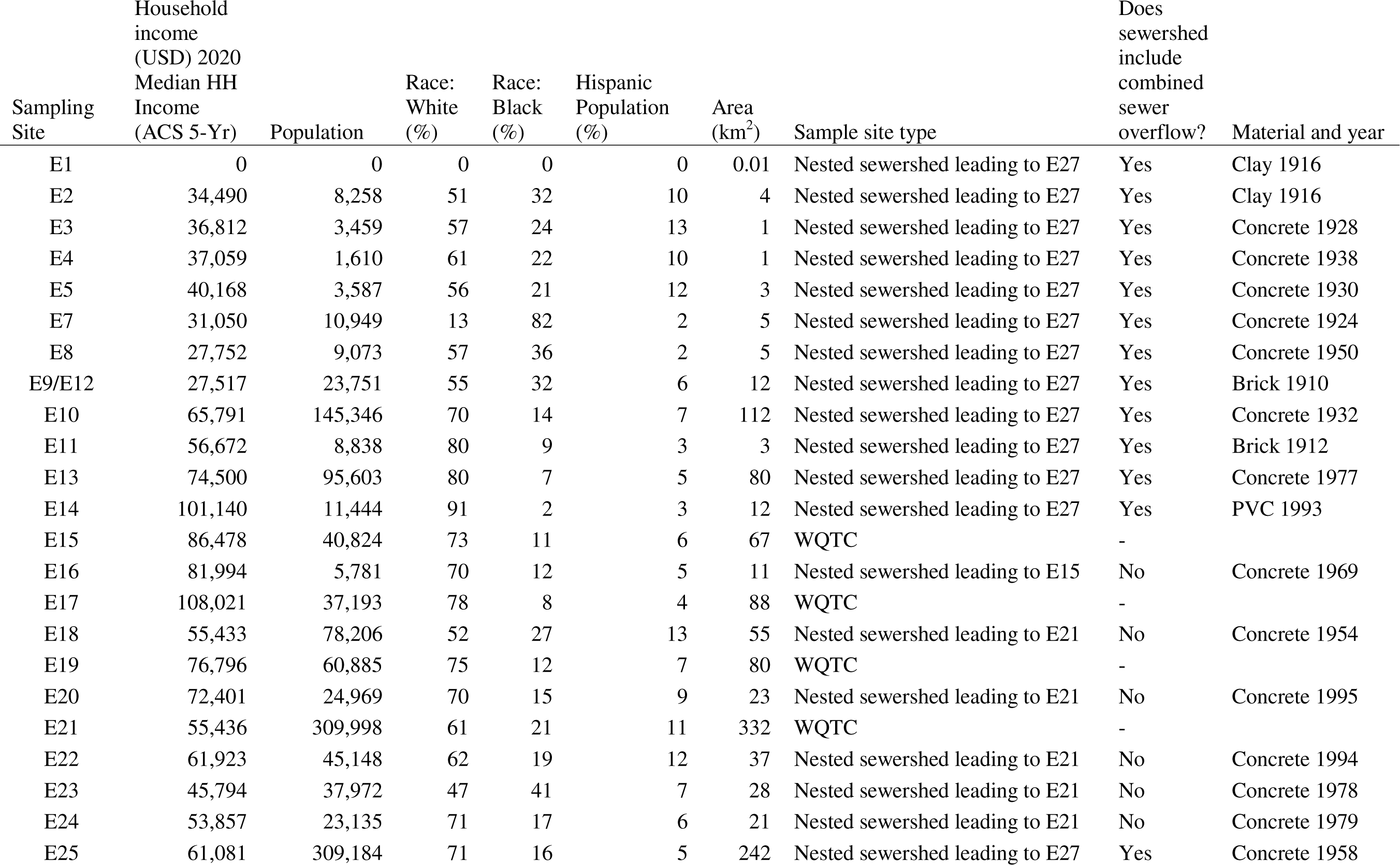
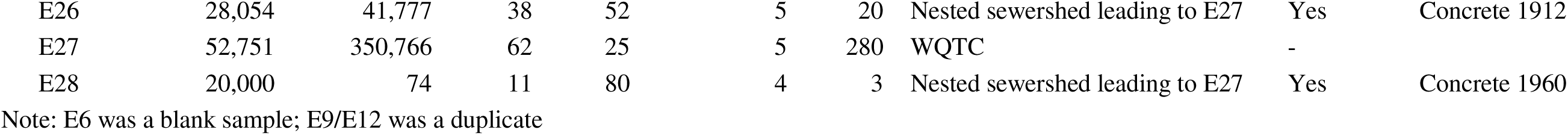
Sample site characteristics, Jefferson County, Kentucky (USA).

For each of the 26 studied metals, sample concentrations with values greater than 1 or 2 standard deviations above the mean were designated areas of concern and hot spots, respectively. Further analysis considered the five-wastewater treatment plant sewersheds and our selection of nested upstream smaller neighborhood/multi-neighborhood scales that flow into the treatment plants for the role of sewershed scale dynamics.

## 3. Results

### 3.1 Places of concern

Nineteen sites had one or more metal results greater than one standard deviation above the mean and were designated areas of concern (Fig. 2) across the county. All 26 studied metals had at least one site of concern (Fig. 3), in total there are 59 instances of exceedance amongst the nineteen sites. The 7 sites with no exceedances were a mix of neighborhood (5) and WQTC (2, sites E27 and E19) sites. Additionally, thirteen of the nineteen sites were of increased concern with levels two standard deviations above the mean and designated ‘hot spots.’ Again, all 26 studied metals had at least one hot spot site, in total there are 37 instances of exceedance amongst the 19 sites. WQTC concentrations tended to be closer to the countywide mean though the 13 sites with no exceedances above two standard deviations were a mix of neighborhood (9) and WQTC (4) sites.

**Fig. 2.**
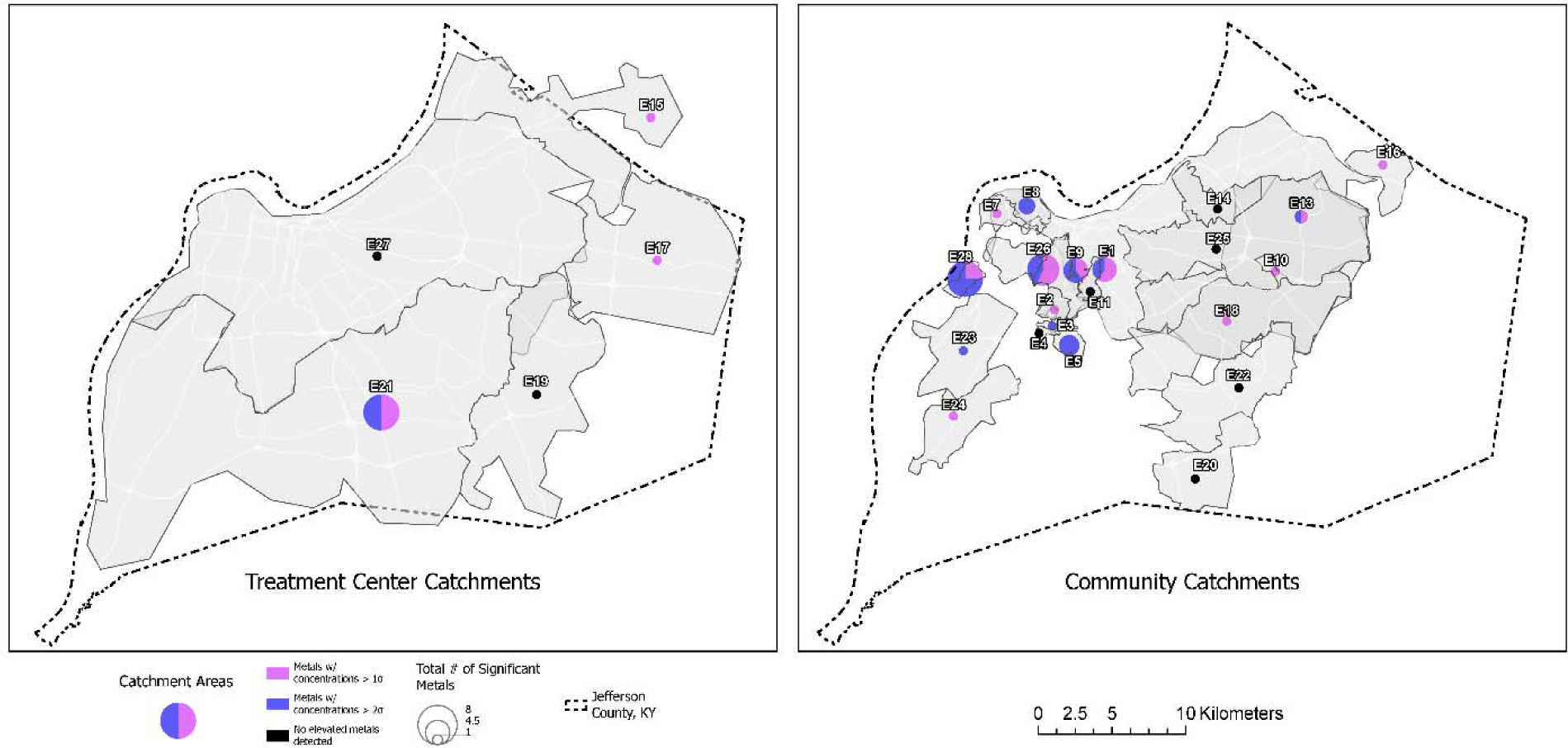
Location of wastewater sampling treatment plants (left) and corresponding sewershed areas (right) which had metal concentrations greater than 1 or 2 standard deviations above the mean, Jefferson County, Kentucky (USA).

**Fig. 3.**
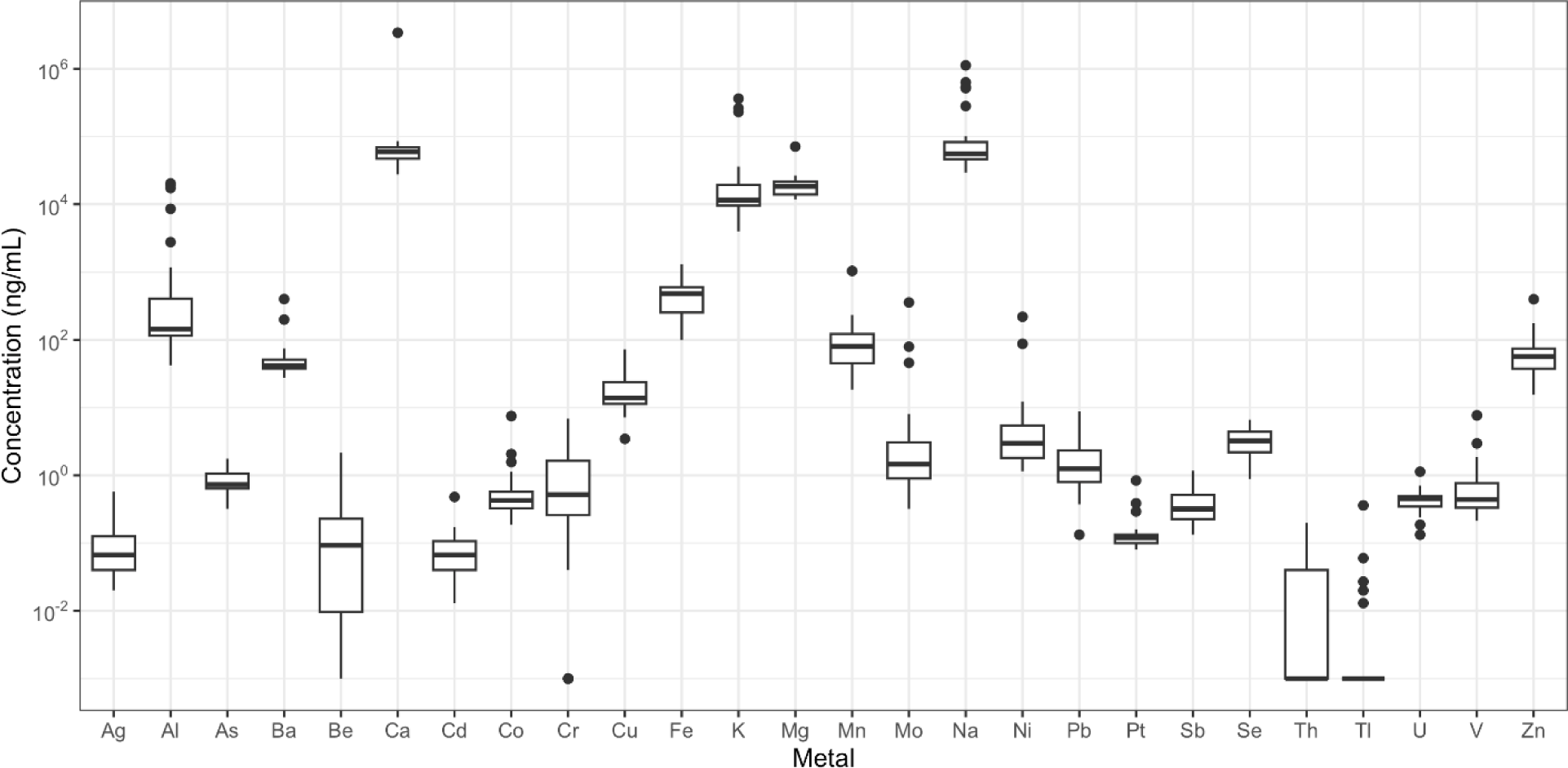
Metal concentration variability for studied sewersheds (N=26), Jefferson County, Kentucky (USA). Metal concentrations are plotted as log concentrations. The box regions represent the first to third quartile, and the black dots represent the outliers. Data were transformed by adding 0.001 to values of 0 or not detected for data distributions.

Both neighborhood sites feeding into the treatment plants and WQTCs had levels of at least one metal 1 or 2 standard deviations above the county-wide mean. Of the five treatment plants, two had a single elevated metal of either Ag (site E17) or Sb (site E15) of only 1 standard deviation above the mean. However, one WQTC (site E21) had levels of Cr, Fe, Cu, Zn, Ag, Sb, Pb, and U greater than one standard deviation above the mean, and of Fe, Cu, Ag, and U were 2 standard deviations above the mean. In comparison, 9 neighborhood sites had a single elevated metal 1 standard deviation above the mean; but it was also common (8/26 sites) for a cluster of two to eight metals at a site. Where there was more than two metal exceedance, no two sites had the same list of metals.

### 3.2 Contributing sewershed areas

Previous work in the study area (Holm et al., 2022) showed for SARS-CoV-2 in wastewater that concentrations across the sewershed area (comparing downstream aggregated treatment plant to nested upstream contributing sewersheds), that the community sites were not different from the respective treatment plants. Variability in metal concentrations is evident in several instances between smaller nested upstream contributing neighborhood sewersheds when measured in the associated downstream treatment plant. The 21 nested upstream contributing neighborhood sewersheds level sites flowed to three WQTCs. Two treatment plants did not have nested sites included in this study due to the small population served.

There are several nested sites interesting cases to detail in regard to variance in arsenic, barium, cadmium, chromium, lead, selenium, and silver which have human health risks (Fig. 4). Lead was observed to be elevated at two sample locations, a neighborhood site (site E23; 8.8 ng/mL) and also the downstream WQTC to which it flows (site E21; 5.48 ng/mL); within this sewershed area the service area user discharge limits allow three times higher than for the other countywide zones. For the remaining sample locations, lead was below 4.2 ng/mL. These results uniquely indicate a high likelihood that the nearly 38,000 residents of sewershed site E23 have lead exposure which is not seen in other areas of the treatment plant zone, or wider county. A similar trend is also observed for Sb at neighborhood Site E16 (1.17 ng/mL) which flows to Site E15 (0.78 ng/mL), where the neighborhood site concentration is higher than the treatment plant.

**Fig. 4.**
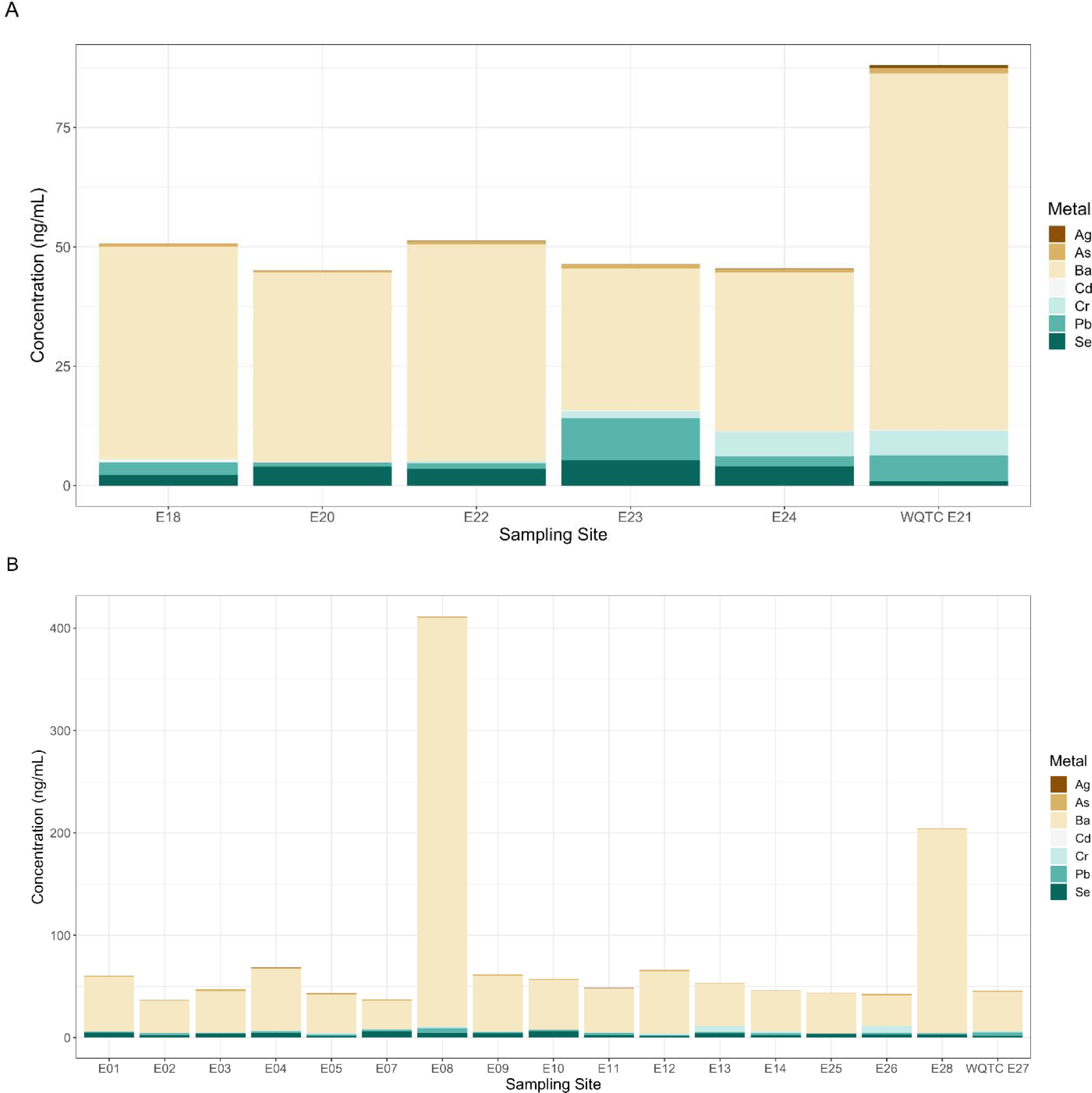
Comparison of metals total concentrations for contributing sites flowing from the upstream locations for arsenic, barium, cadmium, chromium, lead, selenium, and silver versus the water quality treatment center concentration. Derek R. Guthrie Water Quality Treatment Center (Site E21; N = 5 contributing sewersheds) (A) and Morris Forman Water Quality Treatment Center (Site E27; N = 16 contributing sewersheds) (B).

At site 27, which is the largest WQTC in the county and the state, there are no elevated metals. Site 27 has a population of 350,000. But, of the 15 sites that flow downstream to Site E27, 13 sites have exceedances greater than 1 or 2 standard deviations above which are mixed across the countywide and eventually reach the WQTC. Five sites that flow to this treatment plant (Site 27) have elevated levels of a cluster of 4 or more metals; each of these sites has old pipes, in some cases dating back to 1910-1920 with the newest dating back to 1960. That the sewer network for Site 27 is combined sewer whereby stormwater and sewer waste mix in the same pipes may be the cause of additional mixing. The historical infrastructure brings to attention the possibility of pipe leaks and leaching to groundwater as a source of metals contamination and associated rationale for the discrepancy of the results. Site E1 has no households associated with it but is showing elevated metals; a review of the sewersheds revealed industrial land uses including chemical manufacturing and automotive repair but these concentrations of Al (8,580 ng/mL), Ni (87.39 ng/mL), Cu (72.08 ng/mL), Mo (356.17 ng/mL), and Th (0.13 ng/mL) are diluted when they reach the treatment plant.

The exception is observed at treatment plant site E21 which has 8 metals at levels greater than 1 standard deviations above the mean, but only 3 of the neighborhood sites studied are elevated for only one metal each. Because fully encompassing sewersheds were not included, this may just mean we missed coverage and the metals source area was not sampled. But these results indicate a high likelihood that the nearly 310,000 residents of sewershed site E21 have a cluster of metals exposure somewhere within the treatment plant zone.

Of the community sites (21 total), two of the six separate sewer system zone sites had elevated metals (2/6). As well, there were fewer exceedances where the newest pipes were present. Of the three sites with pipes from the 1990s there were no exceedances.

For the seven locations with no exceedances, which may indicate no or comparatively lower metals exposure, they are spread geographically throughout the county and across a range of population size (from 8,800 to 350,0000) and sewershed size (from 280 km^2^ to 3 km^2^). Household income where there are exceedances ranges from the lowest ($20,000) to the highest ($108,000) in our study area, of the two sites with median household income over $100,000, only Site E17 has elevated levels of Ag. Of the two neighborhood sites with the greatest numbers of metal exceedances (7 metals at Site E26 with median household income of $28,054 and 8 metals at site E28 with median household income of $20,000), household income was low. These results disprove that environmental exposure for metals is solely an issue in low-income areas of the county and rather requires countywide surveillance.

### 3.3 Proximity to other environmental sources of pollution

Environmental polluting sources including TRI facilities, brownfield sites, superfund sites, and metal recycling facilities in proximity to our sampling sites could have contributed to the observed metal concentrations (Fig. 5). Most polluting facilities are concentrated in the northwest portion of Jefferson County, which is represented by more low-income households and African American residents. Using arsenic as an example (Fig. 6), countywide concentrations ranged up to 1.76 ng/mL, but the highest levels of metal concentrations are in the northwestern part of the county and the east and southeastern zones were as low as 0.32-0.69 ng/mL.

**Fig. 5.**
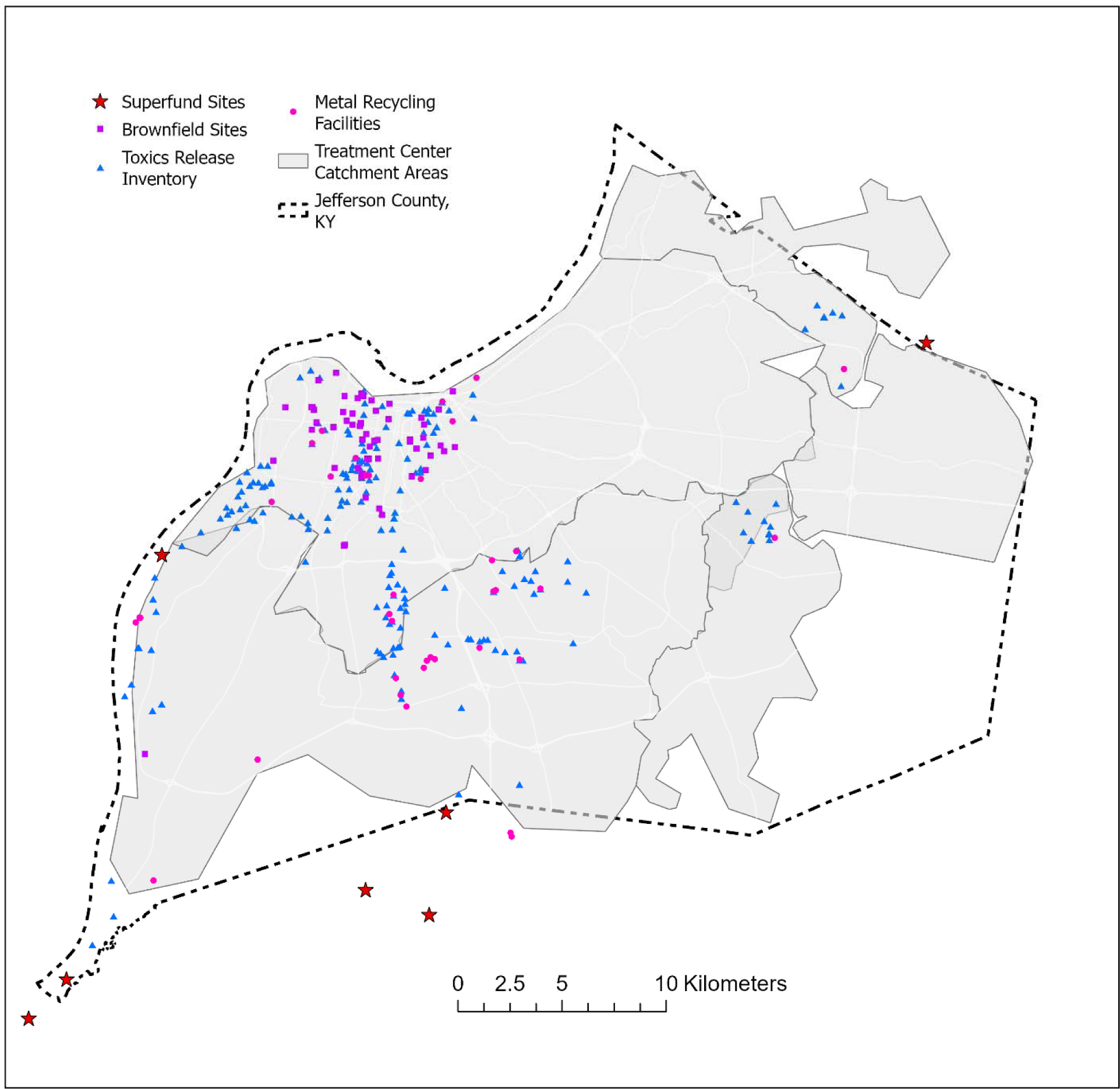
Location of environmental polluting sources including brownfield sites, metal recyclers, superfund sites and toxics release inventory (TRI) facilities, Jefferson County, Kentucky (USA).

**Fig. 6.**
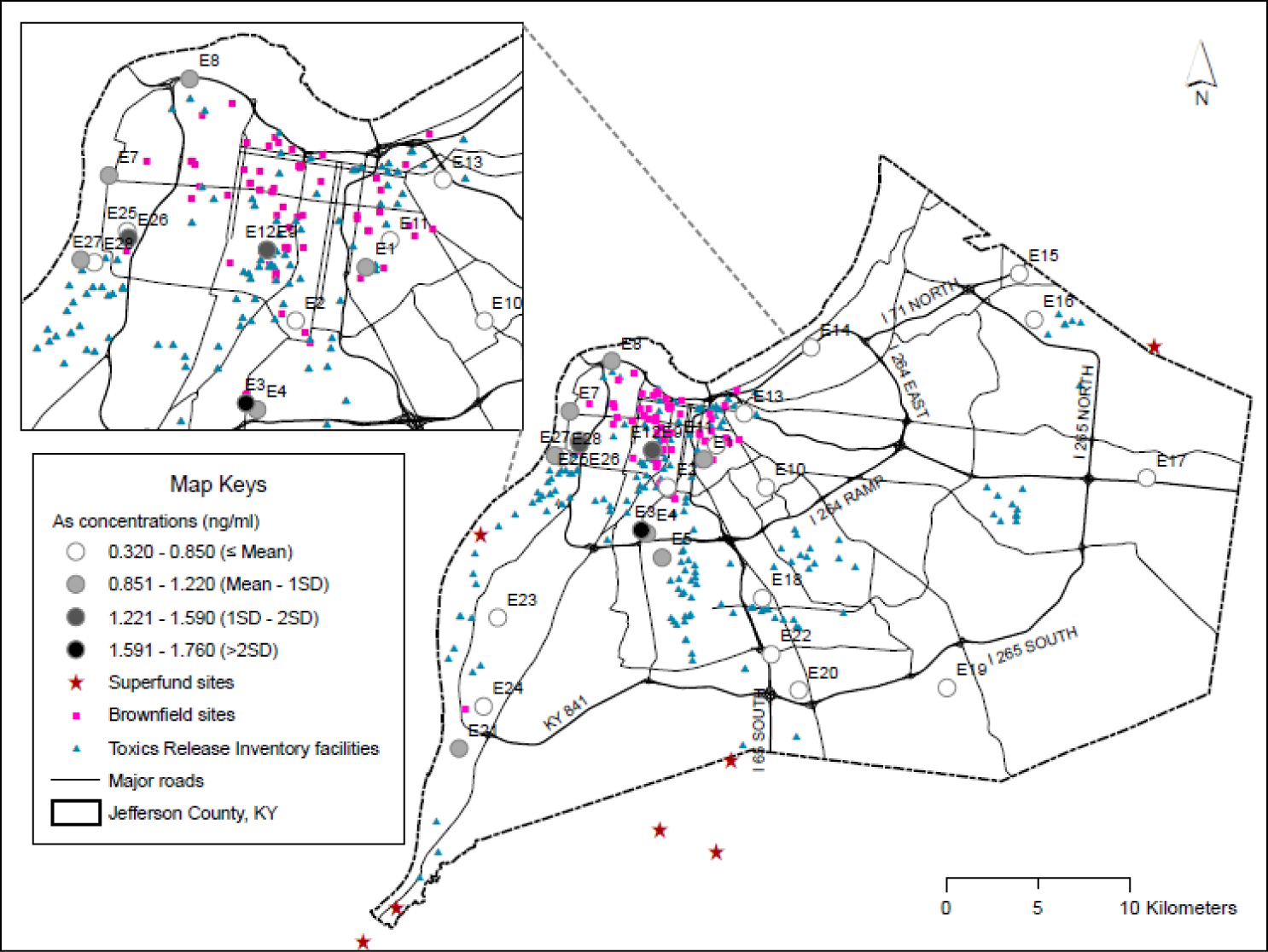
Spatial variations of arsenic concentrations across sampling sites and environmental polluting sources, Jefferson County, Kentucky (USA).

Inclusion or exclusion of environmental polluting sources was not always the only factor, within the sewershed area for Site E17 there are no sources but the site remains with elevated Ag. As well, within the northwest portion of Jefferson County where the majority of these sources are located, a cluster of 6 neighborhood level sites (E9/E12, E13, E8, E7, E11 and E26) we sampled are within the urban core of the county, but only 5 of these sites have elevated metals, and some sites have just one or two elevated metals.

### 3.4 Comparison to Maximum Contaminant Level and service area wastewater discharge permits

In the absence of metal concentrations in wastewater health standards, our results were additionally compared to the limits established by the Safe Drinking Water Act clean drinking water act as an indicator of metal exposure per the enforceable Maximum Contaminant Level (MCL; Environmental Protection Agency, 2009). These were available for 11 of the 26 studied metals. For each site, results were below the MCL (Table 2). For most of the metals (antimony, arsenic, barium, cadmium, chromium, copper, selenium, thallium, and uranium), the maximum metals concentrations in our study were less than 20% of the MCL. However, the maximum concentration of two metals (beryllium and lead) in our study were each about half of the MCL. For beryllium there were 2 samples (2/27) close to half of the MCL; for lead there was only one sample (1/27) close to half of the MCL. EPA (2009) also lists secondary standards for four of the metals assayed (aluminum, iron, manganese, silver, and zinc). Of these, Aluminum, Iron and Manganese were found above the secondary standard for drinking water.

**Table 2.**
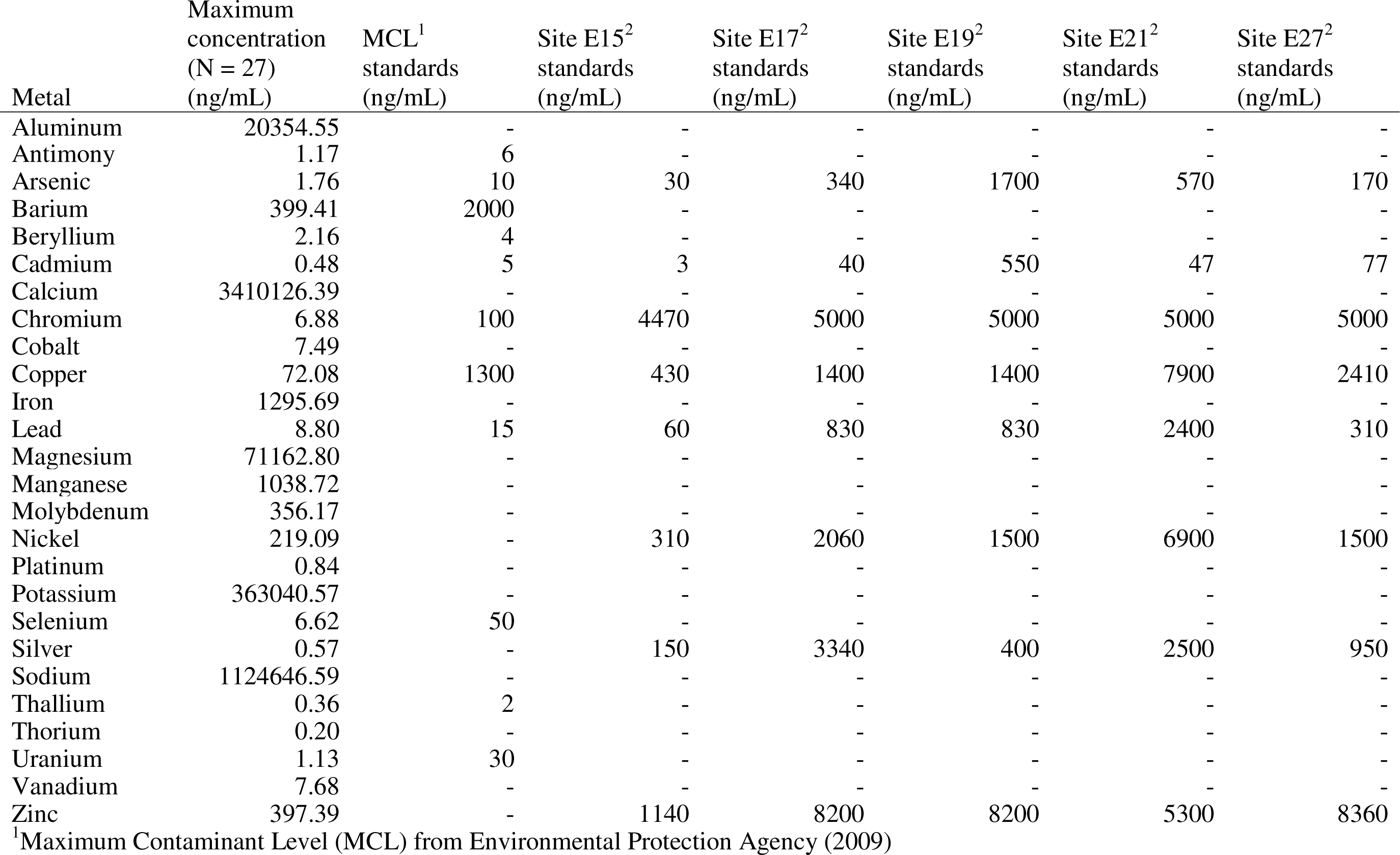
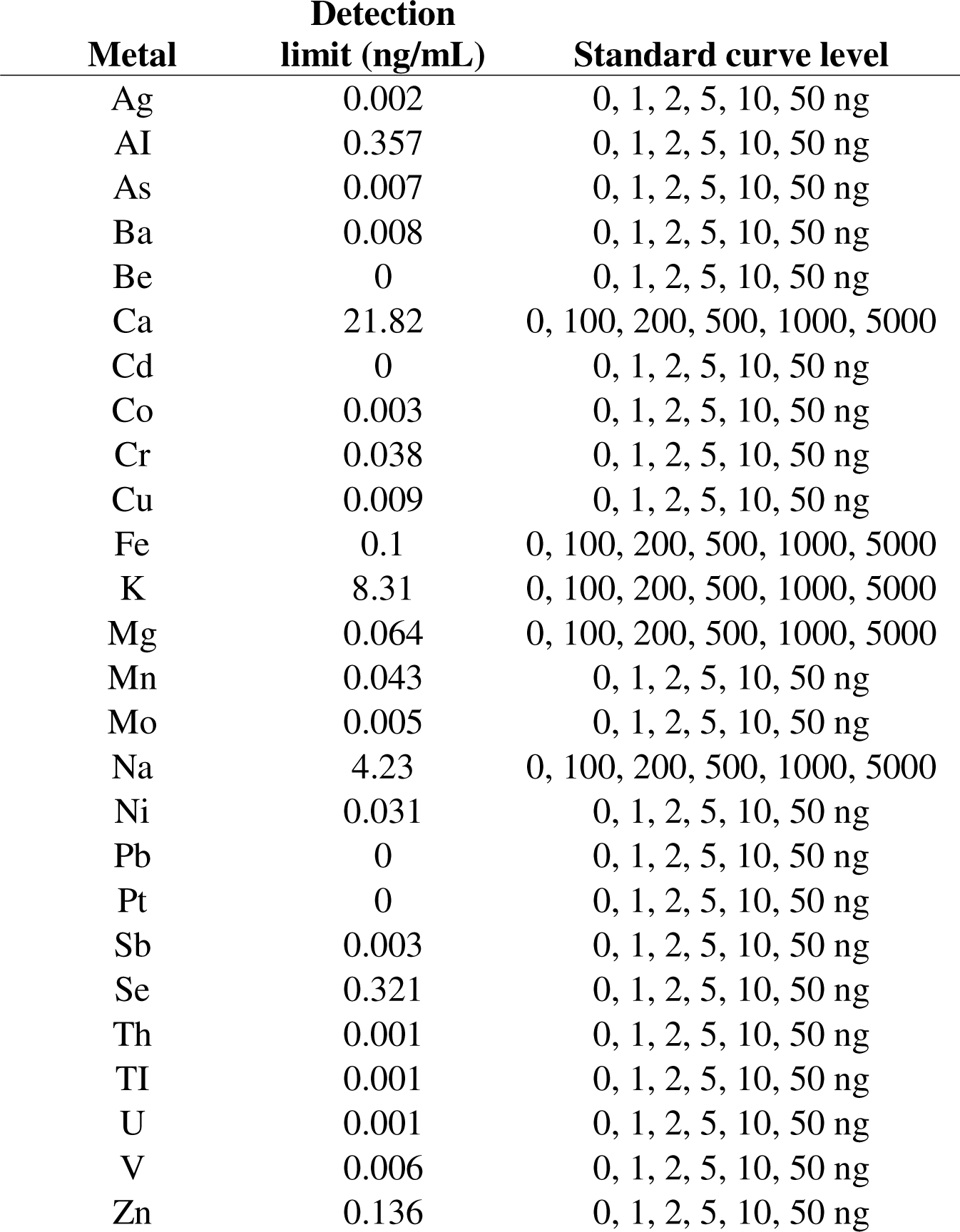
Concentration of studied metals, maximum concentration, Maximum Contaminant Level and five service area user discharge permit standards.

The area user wastewater discharge permit standards (MSD, 2022) are higher concentrations than the MCL standards. The permit limits vary across the county based on the wastewater treatment plant sewershed zone. Industrial input varies from 1 to 30% across the WQTCs. For each studied site, results were below the service area user discharge permit standards (MSD, 2022; Table 2). The maximum metals concentrations in our study were less than 5% of permit standards. Even removing those sites with higher industrial input (∼30%; Site 15 and E15) results in sites that have elevated levels. However, the metals in the samples likely have suffered dilution and do not necessarily reflect the concentrations in any industrial discharge.

### 3.5 Quality control samples

The Environmental Protection Agency (2023a) wastewater sampling procedure recommends equipment rinse blank samples especially in cases of low-level contamination. One equipment rinse blank was collected during field sample collection. Despite that PFAS free water was used, quantified metals were detected for 22 of 26 metals (Table S2).

One site (E9/E12) was sampled at two different times on the same day to test for site variability, approximately one hour apart with two separate field sample collection technicians. The field duplicate sample pair both showed elevated metals, but with different cluster compositions (Table S3). One sample was elevated in Be, Na, Al, K, and Pt, while the second sample was elevated in Be, Na, Al, K, Fe, and As, although all seven metals were detected in both samples. Although a single quality control case, the quantitative variability indicates a requirement for further research attention into temporal dynamics of metals in wastewater. The field duplicate sample site is also uniquely near a neighborhood heavily contaminated with arsenic which was used for manufacture of pesticides and for treating lumber. Much (but not all) of the neighborhood had its topsoil removed and replaced by the EPA. Whether the variability in this site is from temporal differences or a result of the historical contamination needs further investigation.

## 4. Discussion

### 4.1 Community vulnerability

Wastewater can be part of the health equity discussion (Holm et al., 2023b), especially where little or no toxicity data exist in places and times. Wastewater may offer a new environmental matrix to learn how vulnerable residents are exposed to environmental toxicants and guide remediation and prioritize federal and state regulatory requirements, and as needed for emergency surveillance. Certain metals are associated with an increased risk of cancer and other chronic diseases. However, the relationship between metals and cancer is complex, and not all metals are considered carcinogenic. Examples of metals that may increase cancer risk include arsenic, cadmium, nickel, beryllium and chromium (IARC Working Group on the Evaluation of Carcinogenic Risks to Humans, 2012). Because blood and/or urine levels of many of these metals in our study are uncommon, with the exception of childhood blood lead levels, wastewater provides an alternative opportunity to test for community level metals concentration at one time for an aggregate picture of exposome. The gradients of metals observed in this study show that wastewater may provide a unique approach to identifying risk of exposure within a community to guide intervention.

People are exposed to non-essential and essential metals via ingestion, inhalation, and dermal contact. From the initial site of exposure, heavy metals affect nearby tissues and enter the bloodstream to accumulate in sites far from the initial area of exposure. Ingestion though is a major route of exposure. Industrial waste and contamination pollute local air, water and soil affecting heavy metal levels in crops and agricultural animals. Some food crops bioaccumulate specific metals, e.g., grains can bioaccumulate cadmium, and rice can bioaccumulate arsenic.

Thus, when humans ingest food and water affected by this pollution, they ingest heavy metals which can lead to toxicity. Wastewater sampling is beneficial in terms of broad low-cost surveillance of these elements.

The impact of metal toxicity on human health can vary depending on the specific metal involved, the duration and intensity of exposure, and individual factors such as age, overall health, and genetic predispositions, with certain general effects: [1] Metals can target and accumulate in specific organs, leading to damage and dysfunction. For example, lead primarily affects the central nervous system and can impair brain development in children, leading to cognitive and behavioral problems. Lead also targets heme synthesis and can induce anemia. Mercury can damage the nervous system and kidneys, while cadmium primarily targets the kidneys and can cause kidney disease; [2] Metals impact the developing fetus and reproductive health. Exposure to certain metals during pregnancy can result in developmental abnormalities, birth defects, and impaired growth. Some metals, like lead and mercury, can also affect fertility and disrupt the normal functioning of the reproductive system; [3] Many metals have neurotoxic effects and can cause neurological disorders. Symptoms may include cognitive impairment, memory loss, tremors, coordination problems, and sensory disturbances. For instance, mercury poisoning can lead to Minamata disease, a neurological condition characterized by vision and hearing loss, muscle weakness, and in severe cases, coma and death; [4] Some metals, such as lead, arsenic and cadmium, can contribute to cardiovascular diseases. They can promote the development of atherosclerosis (hardening of the arteries), increase blood pressure, disrupt heart rhythm, and lead to heart attacks and strokes; [5] Ingesting or inhaling certain metals can irritate the gastrointestinal tract and cause symptoms like abdominal pain, nausea, vomiting, and diarrhea.

Prolonged exposure may lead to chronic gastrointestinal problems. Metals like arsenic, cadmium, and mercury can accumulate in the liver and kidneys, impairing their normal function. This can result in liver and kidney damage, leading to a range of symptoms such as fatigue, jaundice, fluid retention, and compromised detoxification; [6] Metal toxicity can interfere with the immune system’s ability to defend against infections and diseases resulting in compromised immune responses and making individuals more susceptible to illnesses (Tokar et al., 2013).

### 4.2 Public health policy implications

Due to both exposure situations and dietary intake, we would expect some variation in metals concentrations across the studied areas. The mean values for all the metals analyzed were up to 46 times the levels in tap water samples. The highest wastewater values observed were up to 381.3 times the levels in the tap water samples. Thus, the increases observed in the wastewater must have come from either the environment sources, or excretion from human environmental exposure. With national reporting systems and networks of laboratories in place for infectious disease surveillance, there is an opportunity to extend this framework to other population health risks from the environment. While previous research has shown promise for the surveillance of toxicants including illicit drugs (Banta-Green et al., 2016; Choi et al., 2019; Croft et al., 2020), tobacco metabolites (Choi et al., 2019), and biological agents as weapons (Sinclair et al., 2008), the examination of exposure to pollutants may hold even greater promise when combined with other exposome emphases. Additionally, given the great interest in the social and other environmental determinants of health and matters of environmental justice, better understanding of place-based exposure risk is timely. This work also has proof of concept to support prioritizing resources and expertise for using wastewater monitoring around the EPA’s (2023b) National Enforcement and Compliance Initiatives for 2024-2027 which include air toxics and chemical accidents while supporting environmental justice considerations to protect public health. There is also a role for the American Society for Testing and Materials standards to assess sampling wastewater for metals as part of large-scale natural disasters or chemical post-incident emergency response or impact monitoring.

### 4.3 Future research

There are several areas that the field of comprehensive communitywide exposome surveillance using wastewater should consider. Although the comparability of 24-hour composite and grab samples for exposome investigations using wastewater media has been studied for disease (Kmush et al., 2022), it has not been studied for metals in wastewater which may be more stable. Finer sampling and sampling both over different times of day and different days will be needed to gain a more complete picture and to localize the source areas for the metals. Our studied sites were in both combined and non-combined sewer systems. Sewage in combined sewer systems includes surface runoff in addition to sanitary system. Thus, metals present in these samples may represent runoff from rain on streets and soil in addition to effluent from households and businesses. It will be important to perform more extensive sampling upstream in the sewersheds to determine whether the metals are coming from a specific area. But the contribution of runoff reflects potential environmental exposures in the source areas which can be helpful to capture. Also, we found arsenic in a sample from a site close to an old industrial site that used arsenic for synthesizing pesticides and treating lumber. Soil from the adjoining household yards surrounding this area was removed (although not from all the yards), but the site itself was not remediated. Thus, the arsenic concentration observed suggests wastewater monitoring can be additionally used to detect residual environmental contamination in areas that were heavily contaminated with metals and may not have been adequately remediated. It would also help to look at human toxicology data (from research cohorts) to help interpret wastewater levels. Lastly, case studies are needed in communities if exposome inclusion of metals in wastewater is useful for estimation of waste following large-scale natural disasters or chemical incidents to support emergency response.

## 5. Limitations

The wastewater samples were collected as grab samples, and although a good representation across the county at a specific point in time, composite samples would likely provide a more representative sample. The variability in our field duplicate and equipment rinse blank show the need for quality control samples to evaluate the sampling and handling activities of the investigation; despite not being required for a research investigation, quality control samples following the Environmental Protection Agency (2023b) wastewater sampling procedure, as applicable, is recommended for future work. We cannot make a definitive statement about human exposure without other data sources such as biospecimens from clinical research participants or case data.

## 6. Conclusion

The findings of this foundational research contribute to the existing literature on environmental injustice and shed light on potential linkages between metal exposures measured in wastewater and their concomitant adverse health outcomes after accounting for other social and environmental confounding factors. Wastewater can indirectly capture soil, water, air quality, and human exposure impact for an area with a single sample, widening exposome at a community level. Variability is evident in several instances between smaller nested upstream contributing neighborhood sewersheds when measured in the associated downstream treatment plant; there is a benefit to sampling both scales across a large county. Wastewater provides an opportunity to look at integrated toxicology to complement other toxicology data, looking at where people live and what toxicants need to be focused on to protect the health of people in that area.

## Author contributions

Conceptualization: RHH, TS, JCS; Methodology: LC, RHH, DJB, CHZ, DT, TS, JCS; Formal analysis: LC, RHH, DJB, CHZ, TS, JCS; Writing-original draft preparation: LC, RHH, DJB, CHZ, TS, JCS; Writing-review and editing: LC, RHH, DJB, CHZ, DT, TS, JCS; Supervision: TS, JCS; All the authors have read and agreed to the published version of this manuscript.

## Acknowledgements

We thank the Louisville/Jefferson County Metropolitan Sewer District for their valuable collaboration with the wastewater sample collection. The authors are grateful to Dr. Jason Xu from the Pediatric Research Institute for his excellent assistance for the metal measurements with ICP-MS, and to Mr. J. David Hoetker from the Christina Lee Brown Envirome Institute for his excellent assistance for the wastewater sample delivering and preparation of these samples for the assay.

## Data Availability Statement

Data generated in this study can be found in the article and its supplementary files.

## Supplementary Material

**Table S1.**
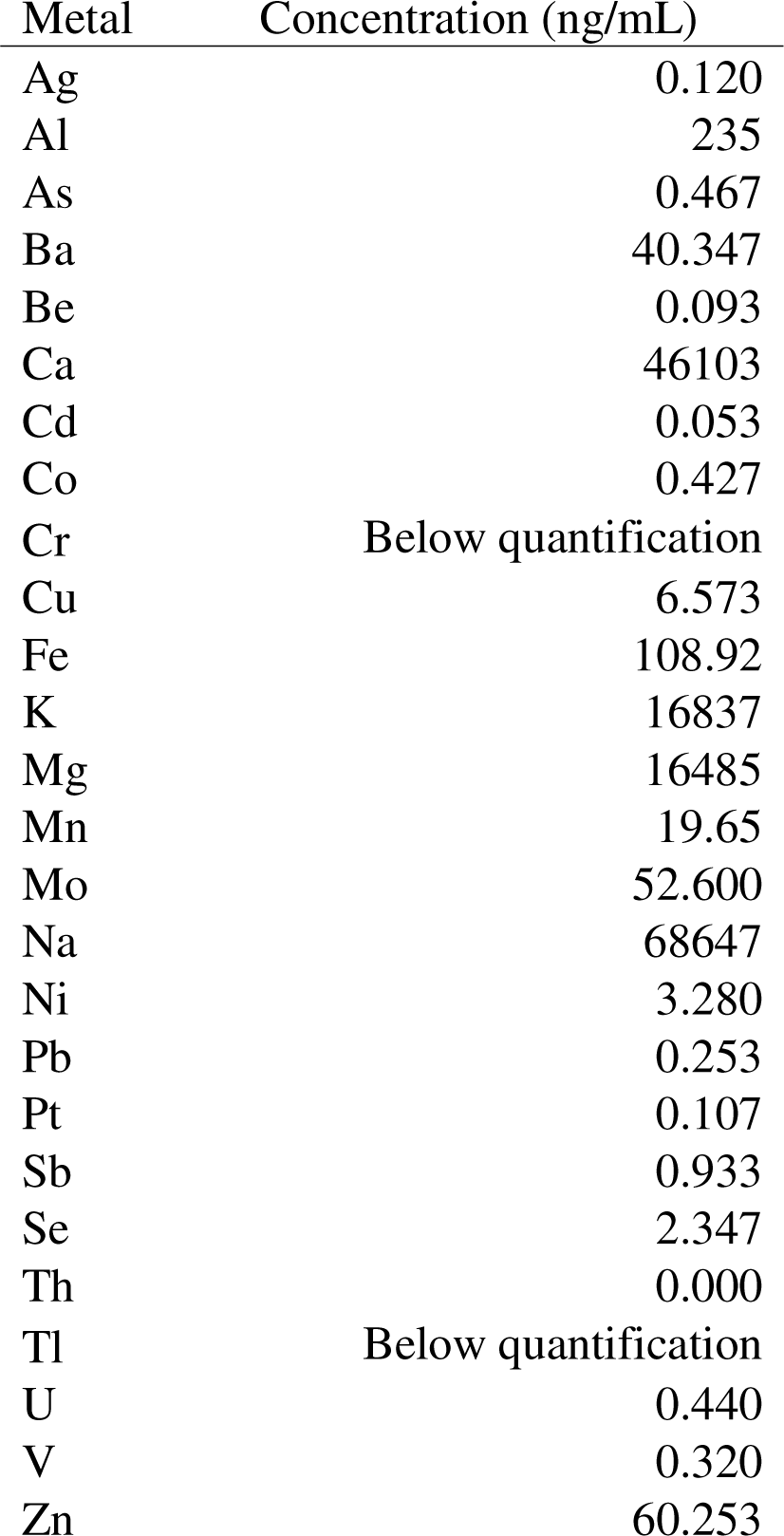
Metals studied by Inductively Coupled Plasma Quadrupole Mass Spectrometer, detection limit, and standard curve levels.

**Table S2.**
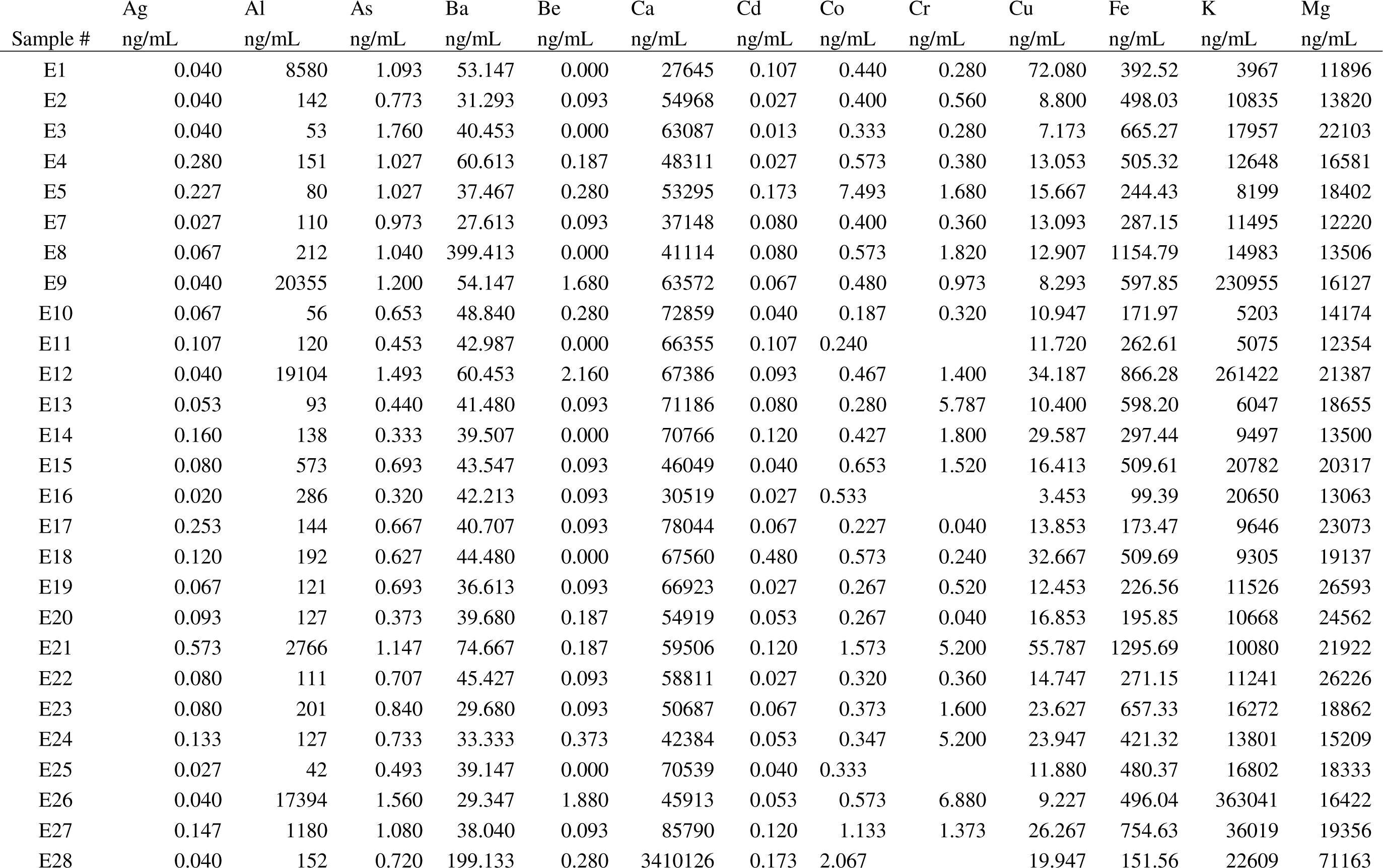
Metal concentrations of equipment rinse blank field sample.

**Table S3.** Mean metal concentrations by site.

| Sample # | Mn<br>ng/mL | Mo<br>ng/mL | Na<br>ng/mL | Ni<br>ng/mL | Pb<br>ng/mL | Pt<br>ng/mL | Sb<br>ng/mL | Se<br>ng/mL | Th<br>ng/mL | Tl<br>ng/mL | U<br>ng/mL | V<br>ng/mL | Zn<br>ng/mL |
| --- | --- | --- | --- | --- | --- | --- | --- | --- | --- | --- | --- | --- | --- |
| E1 | 23.99 | 356.173 | 281538 | 87.387 | 0.707 | 0.120 | 0.187 | 5.000 | 0.133 | 0.000 | 0.133 | 0.507 | 15.453 |
| E2 | 116.47 | 1.667 | 29330 | 4.027 | 1.613 | 0.133 | 0.253 | 2.640 | 0.107 | 0.000 | 0.187 | 0.427 | 38.053 |
| E3 | 201.25 | 1.387 | 49938 | 6.547 | 0.787 | 0.133 | 0.187 | 3.960 | 0.040 |  | 0.240 | 0.213 | 26.347 |
| E4 | 81.57 | 1.160 | 81944 | 6.107 | 1.427 | 0.133 | 0.320 | 4.860 | 0.027 |  | 0.307 | 0.440 | 36.907 |
| E5 | 79.04 | 3.827 | 48540 | 219.093 | 1.027 | 0.120 | 0.533 | 1.760 | 0.200 | 0.360 | 0.347 | 0.547 | 35.173 |
| E7 | 40.85 | 0.813 | 42983 | 1.640 | 1.867 | 0.133 | 0.333 | 6.180 | 0.000 | 0.000 | 0.453 | 0.427 | 28.213 |
| E8 | 233.61 | 1.493 | 46298 | 2.787 | 4.227 | 0.120 | 0.640 | 4.720 | 0.000 |  | 0.493 | 7.680 | 46.013 |
| E9 | 158.79 | 2.267 | 538474 | 1.693 | 0.813 | 0.387 | 0.333 | 4.420 | 0.027 | 0.020 | 0.467 | 1.573 | 78.413 |
| E10 | 82.40 | 0.320 | 39067 | 1.240 | 0.773 | 0.107 | 0.133 | 6.620 | 0.000 |  | 0.453 | 0.280 | 33.547 |
| E11 | 73.99 | 0.360 | 39920 | 1.467 | 2.680 | 0.120 | 0.147 | 2.200 | 0.000 |  | 0.413 | 0.307 | 43.227 |
| E12 | 178.15 | 2.173 | 519013 | 2.787 | 1.253 | 0.293 | 0.507 | 1.320 | 0.013 | 0.000 | 0.493 | 1.880 | 54.600 |
| E13 | 53.69 | 0.613 | 56266 | 2.493 | 1.173 | 0.107 | 0.187 | 4.400 | 0.067 |  | 0.440 | 0.493 | 397.387 |
| E14 | 49.73 | 0.747 | 45989 | 2.320 | 2.027 | 0.107 | 0.200 | 2.200 | 0.040 | 0.060 | 0.467 | 0.440 | 79.733 |
| E15 | 27.91 | 45.787 | 83557 | 4.707 | 3.240 | 0.133 | 0.787 | 2.053 | 0.013 | 0.000 | 0.413 | 0.493 | 115.667 |
| E16 | 18.39 | 79.253 | 78686 | 4.320 | 0.133 | 0.133 | 1.173 | 1.320 |  |  | 0.347 | 0.267 | 57.933 |
| E17 | 22.52 | 1.467 | 49306 | 1.147 | 0.373 | 0.093 | 0.320 | 3.080 | 0.000 |  | 0.467 | 0.347 | 72.520 |
| E18 | 86.19 | 2.187 | 55692 | 3.160 | 2.587 | 0.093 | 0.240 | 2.200 | 0.000 | 0.000 | 0.493 | 0.613 | 70.893 |
| E19 | 22.92 | 1.147 | 56050 | 1.400 | 0.400 | 0.093 | 0.333 | 2.200 | 0.000 | 0.000 | 0.547 | 0.307 | 52.040 |
| E20 | 30.60 | 0.653 | 46599 | 1.907 | 0.880 | 0.120 | 0.373 | 3.960 | 0.000 | 0.000 | 0.600 | 0.320 | 64.333 |
| E21 | 214.87 | 4.813 | 47839 | 12.213 | 5.480 | 0.107 | 0.827 | 0.880 | 0.040 | 0.027 | 1.133 | 1.013 | 175.400 |
| E22 | 68.13 | 1.000 | 43767 | 1.720 | 1.200 | 0.093 | 0.307 | 3.520 | 0.000 | 0.000 | 0.493 | 0.427 | 60.560 |
| E23 | 72.92 | 1.280 | 48357 | 2.733 | 8.800 | 0.080 | 0.520 | 5.280 | 0.027 |  | 0.267 | 0.453 | 75.800 |
| E24 | 84.65 | 0.813 | 56362 | 3.347 | 2.093 | 0.093 | 0.267 | 3.980 | 0.000 |  | 0.293 | 0.267 | 56.613 |
| E25 | 80.11 | 1.000 | 62629 | 4.800 | 0.587 | 0.093 | 0.213 | 3.520 | 0.000 | 0.000 | 0.427 | 0.360 | 27.120 |
| E26 | 128.07 | 2.440 | 630457 | 2.973 | 1.653 | 0.147 | 0.320 | 3.080 | 0.000 | 0.000 | 0.467 | 2.960 | 40.253 |
| E27 | 114.28 | 7.147 | 100994 | 12.120 | 3.320 | 0.160 | 0.480 | 1.760 | 0.053 | 0.000 | 0.427 | 0.987 | 75.787 |
| E28 | 1038.72 | 8.040 | 1124647 | 7.613 | 1.053 | 0.840 | 0.960 | 3.227 | 0.000 | 0.013 | 0.707 | 0.960 | 66.133 |

